# Motor Neuron Disease is not clustered or associated with socioeconomic deprivation in Scotland

**DOI:** 10.1101/2022.02.27.22271535

**Authors:** Yuan-Ting Chang, Patrick K.A. Kearns, Judith Newton, Juan Larraz, Deborah Forbes, Emily Beswick, Danielle Leighton, Gavin Langlands, Robert J. Swingler, Siddharthan Chandran, Suvankar Pal

## Abstract

**Background and Objectives:** Understanding risk factors and spatial variation of risk may provide insights into the pathological mechanism of motor neuron disease (MND). Socioeconomic status (SES) has been hypothesised as a risk factor, but results from previous studies are inconsistent. No spatial epidemiology studies examining geographic clustering of MND have been performed in Scotland to date. We investigated the effect of SES and geography on MND risk in Scotland using the highly curated and nationally representative Scottish MND Register (CARE-MND) that has over 95% case ascertainment.

**Methods:** Patients on the Scottish Clinical Audit Research Evaluation for Motor Neurone Disease platform, with date of diagnosis between 01 Jan 2015 and 28 Feb 2021 were included and geocoded with data zones using their residence at diagnosis. Indirectly standardised incidence ratio (SIR) accounting for age and sex was calculated for each health board. Poisson regression was used to examine the effect of SES on MND risk, adjusted for age and sex of population at risk. Geographic clustering was assessed using global Moran’s Index, Bayesian conditional autoregressive models and SaTScan software.

**Results:** Among 1149 patients included, 1126 (98%) were successfully geocoded. Significantly lower SIR was shown in Highland (0.60, 95% CI 0.44 – 0.80, *p*<0.001). After adjusting for age and sex in Poisson regression, there was no significant effect of SES on MND risk. Moran’s Index indicated a weak but significant spatial autocorrelation (I=0.017, *p*=0.008). However, risk maps using Bayesian modelling showed a relatively homogeneous distribution without distinct patterns across Scotland. Similarly, SaTScan did not detect any significant cluster of high or low risks.

**Discussion:** Our findings support the notion that MND risk is mostly random-distributed and independent of SES in Scotland. Our results are generalizable to other MND populations.

## Introduction

The pathological mechanisms giving rise to motor neuron disease (MND) are not well understood but contributions to an individual’s risk are likely to arise from gene variants, environmental exposures and chance. Around five percent of MND cases cluster in families.^1^ However, multiple studies employing different methodologies indicate that in the non-familial cases, approximately 60% of the variation in MND risk (liability) across cohorts is also heritable (i.e. apparently genetic).^2^ Whilst an increasing number of pathological genetic mutations with incomplete penetrance have been identified, such as the *C9orf72* repeat expansion,^3^ much of this heritability remains unexplained, and in most cases no environmental causal factor is recognized. This leaves a substantial proportion of total variation in risk unexplained and suggests that both genetic and environmental factors are yet to be identified. In general, tobacco use,^4^ exposure to pesticides or heavy metals,^5^ and physical activity^6^ have been implicated as possible risk factors on the basis of inconclusive epidemiological evidence with non-consistent and modest effect sizes. However, it is likely that at least some relevant exposures have yet to be discovered.

The crude incidence rate of MND is estimated to be 1.75 cases per 100,000 person-years (py) worldwide and 2.35 cases per 100,000 py in Western Europe.^7^ Geographical discrepancies in incidence rates might be due to some combination of ascertainment, population age-structure and competing risks as well as real variation in risk.^7^ Consistent with at least some geographic variation in ascertainment being real variation in risk, the *C9orf72* hexanucleotide repeat expansion is found in approximately eight percent of people with sporadic MND of European ancestry, but very infrequently in other populations, reflecting a likely European founder effect.^8^ Whether the geographic distribution of other genetic risk variants contributes to this or whether penetrance of MND risk genes is also modified by spatially-varying environmental risk factors is unknown.

Spatial analysis has shown to be a promising approach in the search for non-ubiquitous genetic and environmental causes of MND. Importantly, regional clusters of MND have been identified around the world.^9–15^ Possible reasons for the geographic variations and clusters of MND have been hypothesized, including exposure to neurotoxic chemicals,^16^ disparities in access to health care services,^17^ latitude,^18^ ethnicity^17^ and genetic risk.^17^ The British Isles makes an interesting case study for spatial analysis because the population experiences a wide range of living environments and excellent tools of human geography (accurate age-specific small-area population denominators) are publicly available and maintained. While studies of spatial epidemiology of MND have been conducted in Ireland^19^ and parts of England,^15,20^ which found two low-risk clusters in Ireland^19^ and some high-risk clusters in England,^15,20^ including one statistically significant in London^15^, no risk mapping or cluster analysis has yet been performed in Scotland.

In addition to geographic location of residence, SES is a possible risk factor of interest as it is a surrogate for a wide composite of environmental exposures. In addition to providing aetiological insights, an understanding of spatial MND risk based on SES is also useful in resource allocation, care planning and potentially for clinical trial design. However, only a few studies have investigated the association between SES and MND risk, which revealed positive,^21–23^ negative^24,25^ and non-significant associations.^26–29^ Differences between study conclusions can be explained by use of different indicators of SES, exposure windows and study designs. Only one case-control study^28^ published in 1993 examined the indicators of SES in childhood and their association with MND specifically in the Scottish population. A recent Scottish study^30^ looked at the crude number of cases in SES classes and found no difference.

Scotland is suitable and advantageous for studying MND spatial epidemiology due to the national MND register. The register, established in 1989, was the first population-based MND register and rebooted in 2015 as Clinical Audit Research and Evaluation of MND (CARE-MND).^31^ With exceptionally high rates of case ascertainment (>95%) and extensive clinical phenotyping, the register has enabled researchers to investigate several important questions about MND.^32^ Of particular note, Leighton et al.^30^ reported analysis from the Scottish National MND register, finding that the standardised incidence in Scotland was 67% higher even than the Northern European estimates in 2015-2016.^30^ MND specialist nurses are ideally placed working in localised areas. A focus group with the MND Specialist nurses was established to investigate personal anecdotes of community nursing over two decades. This group identified an area with a perceived high number of cases, as well as two conjugal pairs of MND which have occurred in the decade prior to our study.^33^ The area is centered on the small town of Tullibody (*n*=8580 in 2016) and neighbouring towns, historically a coal-mining community.

In this study, we used data from the Scottish MND register between 2016-2021 to examine for evidence of clustering, non-random spatial distribution of incident cases and associations with socio-economic measures of deprivation. In addition, we performed a focus test based on prior suspicion of an area identified by MND specialist nurses as a possible centre of high-incidence in the two decades prior to the study period.

## Materials and Methods

### Data sources and geocoding

Incident case data were obtained from the CARE-MND.^31^ Inclusion criteria were participants with date at diagnosis of any type of MND between 1 January 2015 and 28 February 2021. Capture-recapture analysis suggests that CARE-MND ascertains in excess of 99% of all MND cases over this period in Scotland.^30^ Those without residential information at diagnosis (*n*=23, 2%) were excluded from the spatial analyses. Ethical approvals for the CARE-MND were granted by the Scotland A Research Ethics Committee 15/SS/0126.

The geographic boundaries for this study were data zones (DZ) from 2011 UK census. In total 6,976 DZs were originally generated based on population threshold, where every DZ contained around 500-1000 people. DZ boundaries are designed based on various local boundaries (e.g. school catchment areas) and after local consultation to ensure they reflect real communities and population estimates are updated yearly and boundaries refined every decade. DZs are the ideal unit of measurement for this study since they are small (mean population 800 persons) and stable enough to allow us to detect any cluster on a practical scale which further investigation could be proceeded. Patients were assigned to a DZ where their postcode of residence fell within it at the time of diagnosis.

The population data were DZ-based mid-year estimates published annually (https://www.nrscotland.gov.uk/). Population in 2020 and 2021 were not available at the time of data extraction and thus were estimated by the interpolation methods with data in 2019 and 2018.

### Variables of interests – socioeconomic status, overcrowding rate and urban/rural

The measures of socioeconomic status are measures from the Scottish Index of Multiple Deprivation (SIMD) 2016,^34^ indicating relative deprivation across 6,976 DZs. SIMD is a composite score derived from several domains, including income and health. Quintiles of overall SIMD ranking were treated as categorical variables in our study. Overcrowding rate is one of the indicators of SIMD. A house is overcrowded when “the number of people sleeping in a home exceeds the room standard or the space standard”.^35^ To single out the effect of overcrowding rate, which has been suggested by previous study,^24^ it was also examined as an independent variable. Urban and rural areas are classified based on population threshold. Urban areas are settlements with populations of 10,000 or more whereas rural areas are settlements with populations fewer than 3,000. Towns are those with populations between 3,000 and 10,000.^36^ The information above was available on the Scottish Government website (https://www.gov.scot/).

### Incidence rate, cumulative incidence, standardization and Poisson regression

Nationwide and sex-stratified incidence rates per 100,000 py were calculated. Age-adjusted incidence rates were also obtained by direct standardisation to the European Standard Population.^37^ Cumulative incidence stratified on sex was calculated by the method previously described.^38^ Logistic growth curve was fitted separately for cumulative incidence stratified on sex. Estimated lifetime risk of MND was modelled by the fitted logistic curves and calculated using life expectancy at birth in the UK (Male: 79 years; Female 83 years).^39^ Upper asymptotes of the fitted curves were reported and interpreted as estimated cumulative risk of MND. Confidence intervals for incidence rates and cumulative incidence were obtained via exact method^40^ and 10,000 bootstrap samples, respectively.

To assess the regional difference in the incidence rates, indirect standardisation was employed. Nationwide incidence rates stratified on age and sex were multiplied by the population structure in each region of interest to obtain the expected cases. Standardised incidence ratio (SIR) of each region was then calculated as observed cases divided by expected cases. Exact Poisson confidence intervals were constructed for SIRs.

Poisson regression was implemented to examine the effects of SIMD, overcrowding rate, and urban/rural classification. Observed cases in each DZ were modelled as discrete Poisson outcomes. Two models were fitted: one with the natural logarithm of the expected count based on the population at risk in each sex and age category as an offset (adjusting for nuisance variables of population age-sex-structure); and the other one with the natural logarithm of the DZ-based population in total. Coefficients were exponentiated and reported as relative risks (RRs).

### Global autocorrelation, risk mapping, and cluster detection

To assess for spatially-autocorrelated unmeasured confounding, global Moran’s I and Bayesian smoothing methods were implemented. The R ‘spdep’ package^41^ was used to create contiguity neighbour matrix of DZs. Because there are 10 DZs on Scottish islands without neighbours, artificial neighbours were created based on the locations of bridges and ferry crossings. Bayesian conditional autoregression with Leroux model^42^ was implemented using the R ‘CARBayes’ package^43^ modelling for global and local random effects. Models were fitted using the two offsets aforementioned and 300,000 runs were performed with 100,000 burn-in samples and thinning interval of 20. Posterior RRs of Bayesian models were used to map smoothed risk.

SaTScan software (v 9.6) was used to detect spatial clusters of MND risk in this study.^44,45^ Scanning was performed separately for spatial clusters of higher or lower than expected rates, adjusting the expected number of cases for the age and sex of the underlying population. The maximum circular window size was set to be 50% of the population at risk. Although there is no established hypothesis of MND cluster size, the maximum reporting size of clusters were set to be 30km based on the findings of previous literature of relatively small, local clusters as well as to enhance interpretability and practical actions (outbreak investigation) upon detection of clusters. Details were described in eMethod. In sensitivity analysis, we removed the restriction of maximum reporting size and scanned with elliptic windows.

Subgroup analyses of cluster scan and Bayesian smoothing were performed stratifying on gender, age at diagnosis with a cut-off at 55 years and classification of MND (including ALS, progressive bulbar palsy, and MND with frontotemporal dementia; primary lateral sclerosis and progressive muscular atrophy were not compared due to limited number of cases). The rationale for an age-stratified examination was that cases with a genetic predisposition would be likely to be over represented in the strata diagnosed at younger ages.^46^ A focused cluster test was also performed with the centre set at Tullibody (the small town observed to have had an unusual abundance of cases in the period before that covered by our study) with circular and elliptical scanning windows.

Analyses except those using SaTScan were conduct via R software (v 4.0.2) and R Studio V. 1.2.5042.^47^

## Data Availability

Data from the CARE-MND platform in this study are not publicly available due to patient identifiable information. Qualified investigators may request access to anonymized data for a reasonable purpose.

## Results

### Demographics, incidence rate and cumulative incidence

Overall 1149 individuals were diagnosed during the study period. 1126 (98%) were successfully geocoded and included in spatial analysis. 690 (60.1%) were males. Mean age at diagnosis was 67.6 (SD 11.5) years. 86 individuals (7.5%) had a family history of MND. The majority had ALS (76.9%). Other types of MND included progressive bulbar palsy (9.9%), primary lateral sclerosis (2.9%), primary muscular atrophy (2.2%), and MND with frontal temporal dementia (6.6%).

Crude and directly standardised IRs were 3.43 (95% CI 3.23 – 3.63) and 3.48 (95% CI 3.28 – 3.69) per 100,000 py, respectively. Higher IRs were observed in male compared to female (Crude IRs Male 4.23, 95% CI 3.92 – 4.56; Female 2.66 95 % CI 2.43 – 2.93, per 100,000 py. Age-standardised IRs Male 4.52, 95% CI 4.18 – 4.88; Female 2.59 95 % CI 2.37 – 2.85, per 100,000 py). IRs increased with age, peaked at the age group 75-79 years and decreased subsequently in both genders (Figure. 1A). Gender-specific cumulative incidence calculated by the data was displayed in Figure. 1B. Estimated lifetime risk of MND modelled by the fitted logistic curves were 3.92 (95% CI 3.69 – 4.16) and 2.61 (95% CI 2.42 – 2.83) per 1000 persons in male and female, respectively. The asymptote of the fitted curve is at 6.02 (95% CI 5.93 – 6.13) and 3.15 (95% CI 3.09 – 3.22) per 1000 persons for males and females respectively.

**Figure 1.**
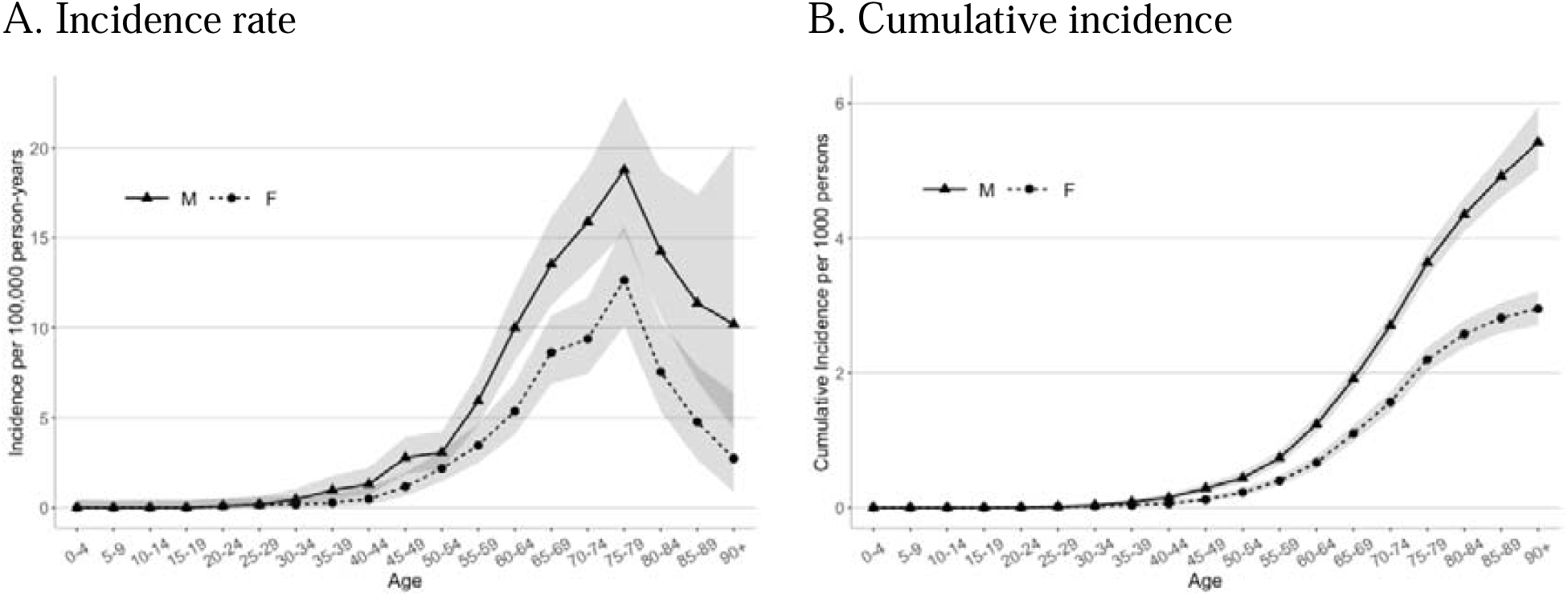
Observed incidence rate (A) and estimated cumulative incidence (B) of motor neurone disease in Scotland. Observed incidence rate (A) and estimated cumulative incidence (B) of motor neurone disease in Scotland (Grey bands indicate 95% confidence intervals.)

Geographic variations observed IRs across health boards, ranging from 2.37 to 5.87 per 100,000 py (Table 1). Whereas 95% CI of SIRs in most health boards covered 1, SIR in Dumfries and Galloway (1.38, 95% CI 1.04 – 1.80) showed evidence of excess observed cases, albeit non-significant. Conversely, SIRs in Highland (0.60, 95% CI 0.44 – 0.80, Bonferroni-adjusted *p*<0.001) and Lanarkshire (0.81, 95% CI 0.66 – 0.97) indicated that these two regions had lower observed cases than expected. Nevertheless, only SIR in Highland was statistically significant after conservative adjustment for multiple testing.

**Table 1.**
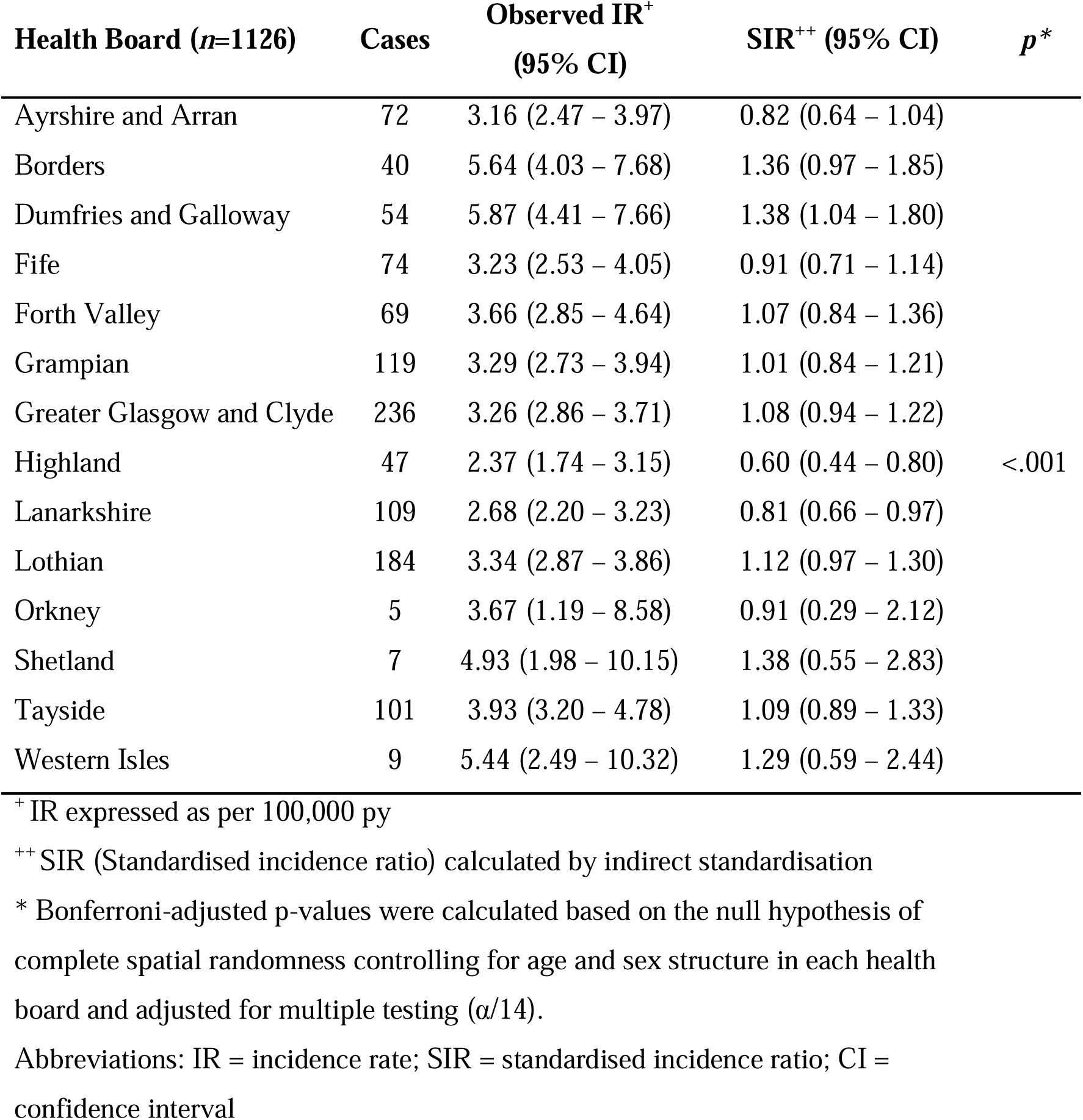
Observed Incidence Rates and Standardised Incidence Ratio of MND across Heath Boards in Scotland.

### Poisson regression

When using crude incidence rates as outcomes, areas that were among the two least deprived categories (SIMD 5^th^ and 4^th^ quintile) had significantly higher MND risks compared to areas most deprived (SIMD 1^st^ quintile) (SIMD 5^th^ quintile: RR=1.28, 95% CI=1.06 – 1.54, *p*=0.010; SIMD 4^th^ quintile: RR=1.24, 95% CI=1.02 – 1.49, *p*=0.032). Increases in household overcrowding rate were also independently associated with significant lower MND risk (RR per 1% increase in overcrowding rate=0.98, 95% CI=0.97 – 0.98, *p*<0.001). Moreover, urban areas were also associated with lower MND risk compared to rural areas (RR=0.81, 95% CI=0.70 – 0.95, *p*=0.007). However, after adjusting for age and sex structure of the population at risk in each small area, these effects were all markedly attenuated, and none were significant (Table 2).

**Table 2.**
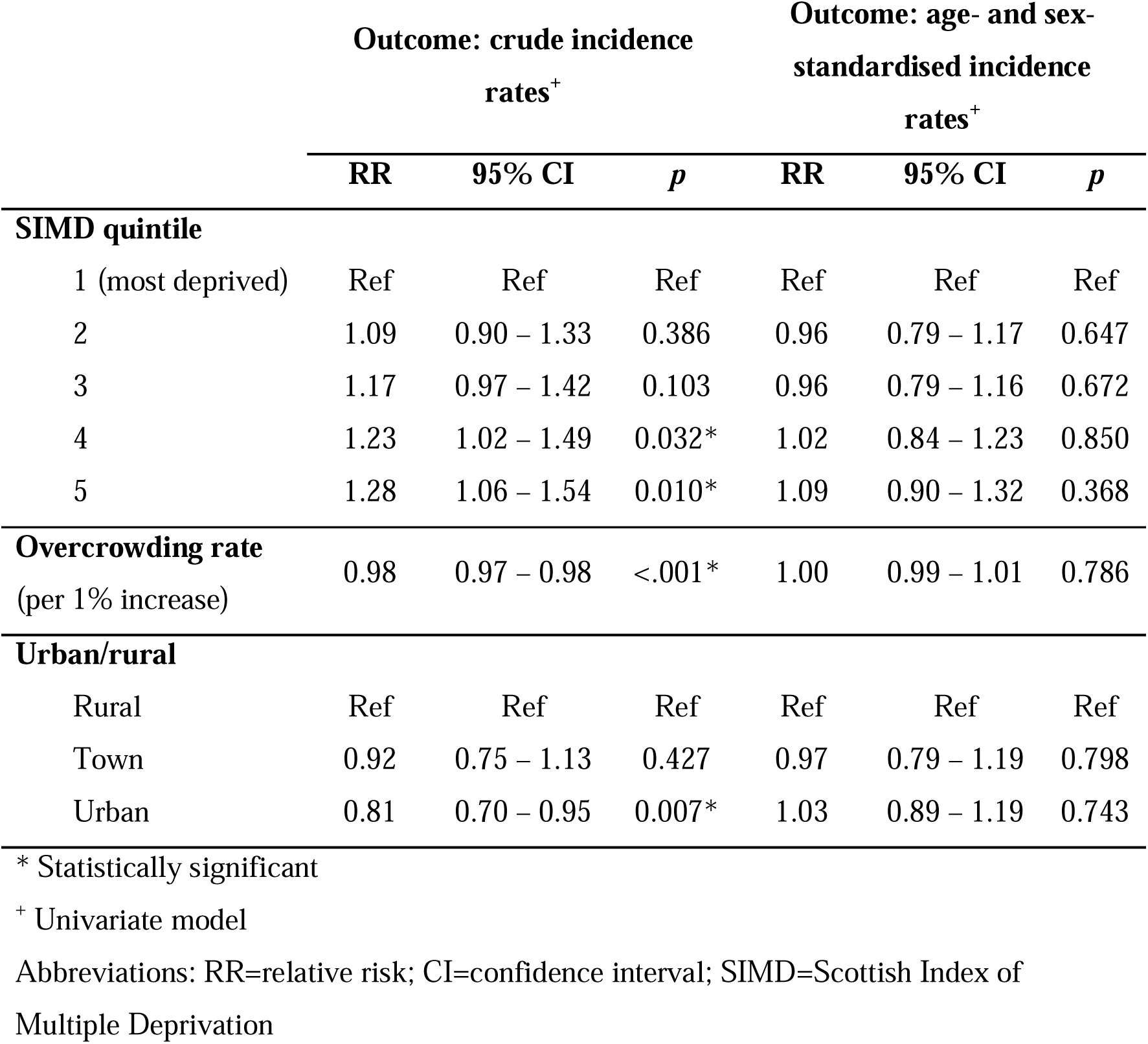
Relative Risk of Motor Neuron Disease, by SIMD Quintile, Overcrowding Rate, and Urban/rural Classification (*n*=1126)

### Risk mapping and cluster analysis

Global Moran’s I showed a weak but significant spatial autocorrelation after adjusting for age and sex of the underlying population in each area (I=0.017, *p*=0.008). Posterior RRs after Bayesian smoothing modelling were mapped in Figure. 2. Whether using crude or age-sex-standardised incidence rates as outcomes, smoothed RRs were relatively homogeneous across Scotland, ranging from 0.98 to 1.03. However, the risk pattern showed more heterogeneity on a local scale in the crude risk map compared to the adjusted one. Smoothed risk maps stratified on gender, age at diagnosis and types of MND also showed homogeneous risks (smoothed RRs ranging from 0.99 to 1.03) with subtle geographical variations across Scotland. (eFigure. 1-3)

**Figure 2.**
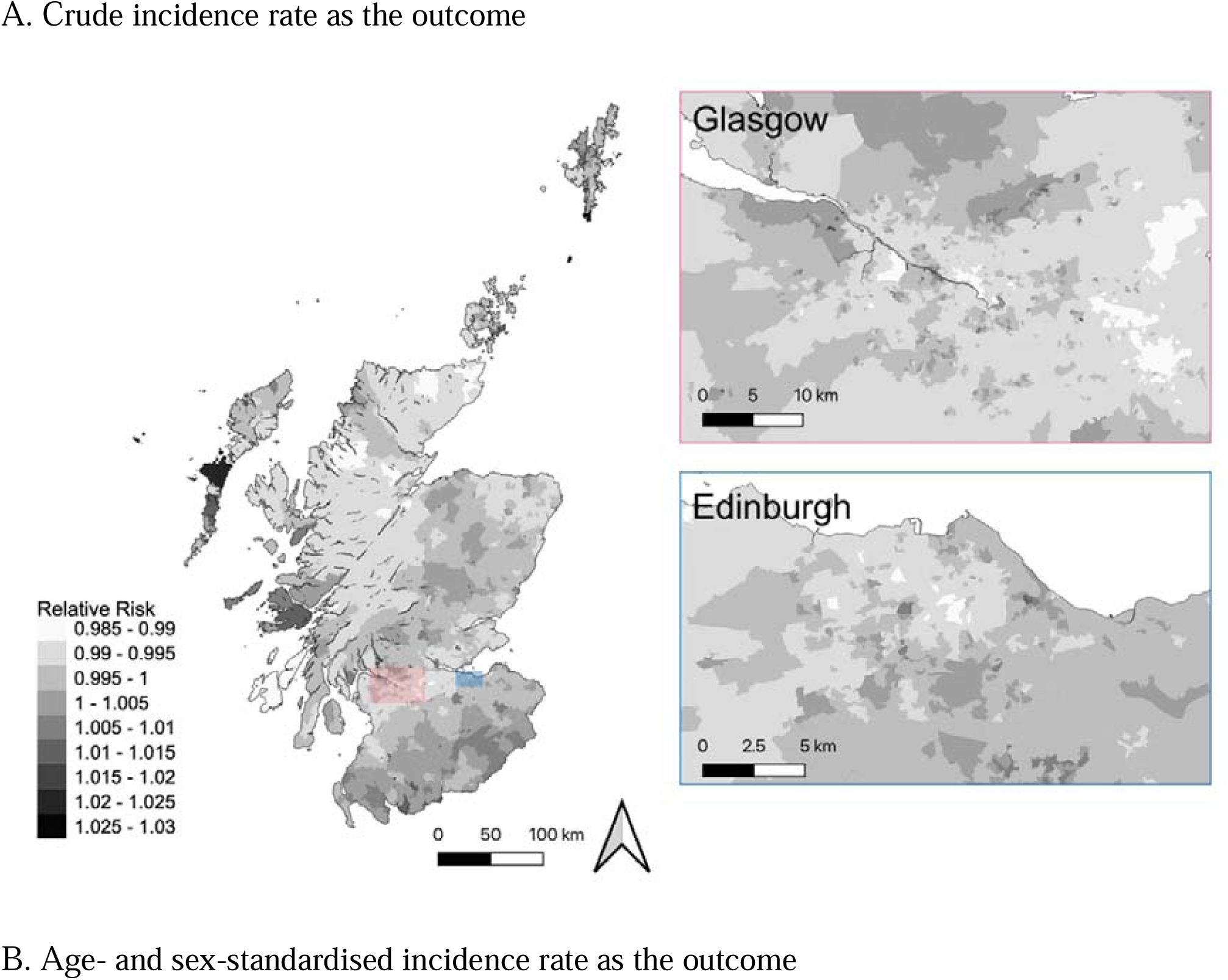
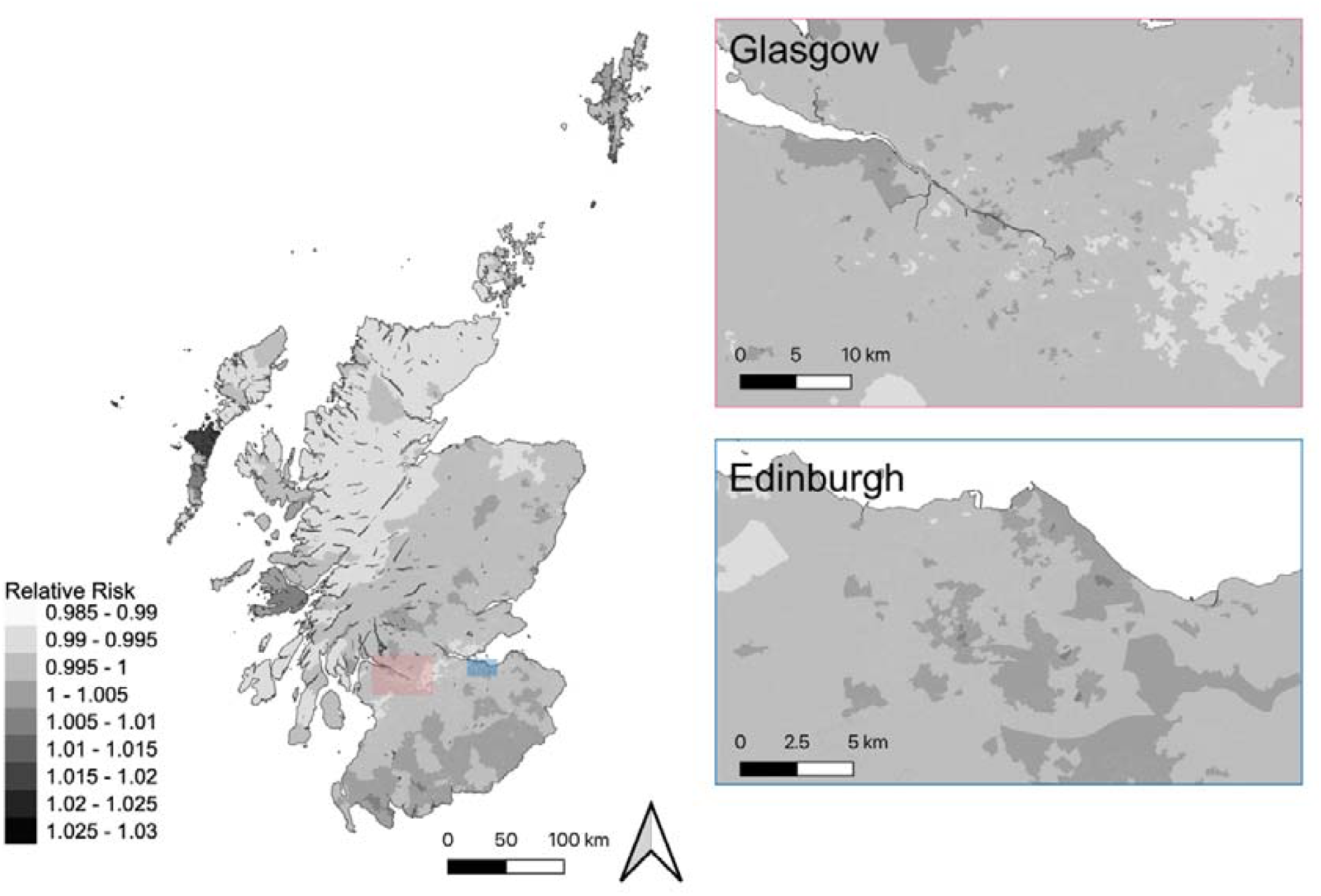
Mapping Bayesian Posterior Relative Risks of Motor Neuron Disease in Scotland.

SaTScan identified three spatial clusters with high incidence rates. (Figure. 3) Two of them were around the city of Glasgow while the other one was in Dundee. However, none reached statistical significance. We reviewed the cases within possible clusters, investigated prospectively collected epidemiological data recorded in the CARE-MND database on occupation and family history, and undertook semi-structured interviews with the local MND specialist nurses who cared for these patients regarding these risk factors and any other factors. No epidemiological links were identified to link the cases. Details of the review were summarised in eAppendix. Characteristic of the nature of MND as a rare disease, unsmoothed RRs across Scotland varied widely even between neighbouring small areas due to the sparseness of cases (eFigure. 4). Scanning for spatial clusters with low rates did not show any statistically significant “cold spots” with low incidence either (eFigure. 5). Sensitivity analyses where maximum window reporting size was not limited (eFigure. 6) and where elliptic windows (eFigure. 7) were implemented also showed consistent findings, failing to detect statistically significant clustering. When we specifically scanned clusters centre Tullibody, where a high number and two conjugal pairs of cases were identified by MND Specialist nurses, the focused cluster test centre Tullibody did identify a possible cluster, but this did not reach the arbitrary significance level (radius=15.54 km, *p*=0.391).

**Figure 3.**
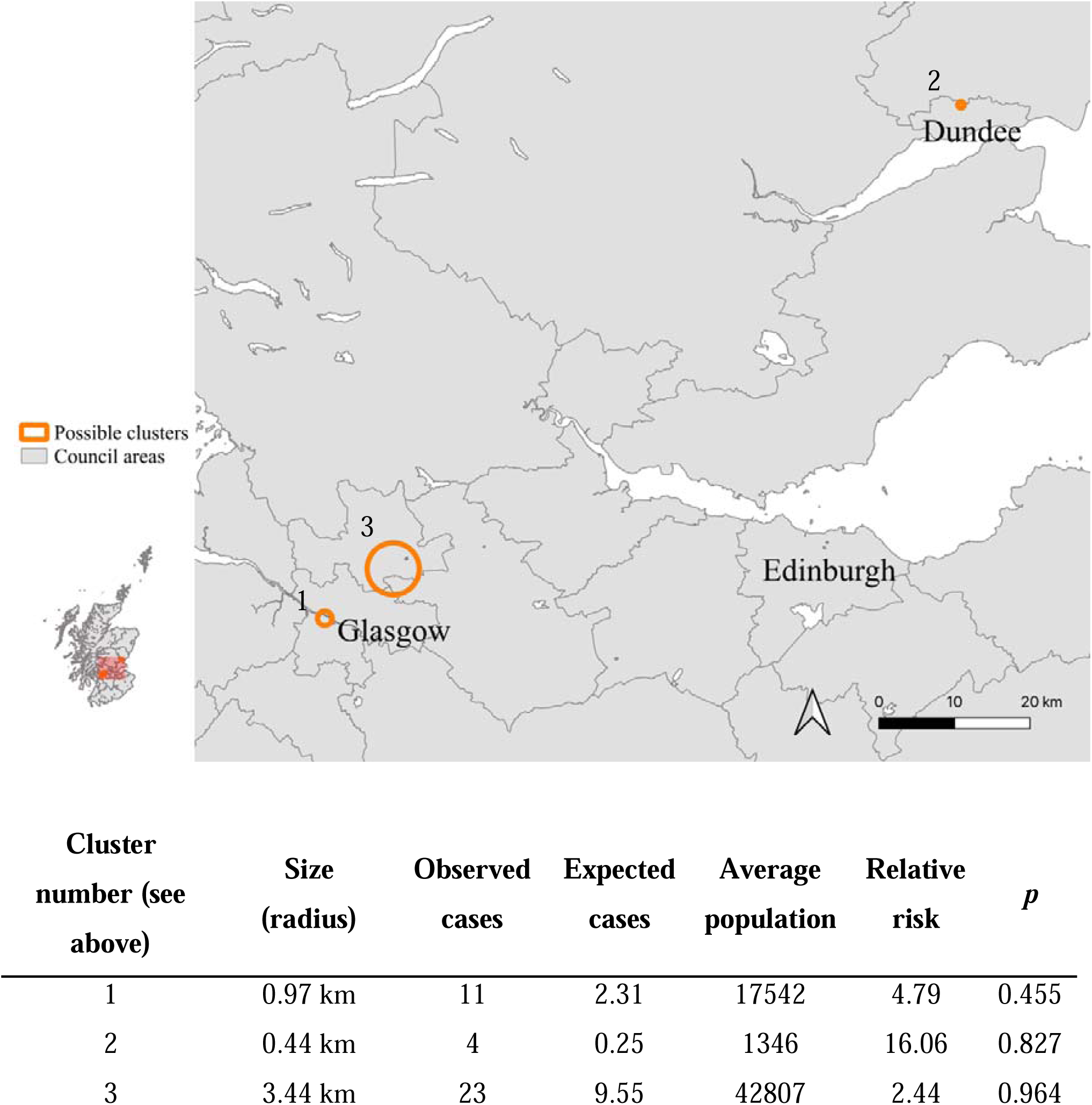
Possible Clusters with High Rates of Motor Neuron Disease in Scotland without a priori suspicion.

## Discussion

In this study we used population-based data from the Scottish national CARE-MND register to describe incidence rates of MND and investigate the effects of SES and spatial location on the risk of MND in Scotland. Despite apparent spatial differences between areas when examining crude rates, our results suggested that after careful adjustment for important confounders in the sex- and age-structure of underlying population in the Poisson regression models, the risks of MND were not significantly different between different levels of SES, overcrowding rates, urban and rural areas. This adjustment was only possible due to the availability of accurate age- and sex-stratified population denominator data in Scotland that is available for small areas and may caution against uncritical overinterpretation of apparent associations in other settings where adjustment for these important factors is limited by data availability. Although some variations of incidence rates were found across health boards and there was some indication that, as a whole, cases were not entirely randomly distributed, no statistically significant clusters with high rates were detected via SaTScan. Bayesian smoothed RR maps adjusting for the sex- and age-structure of the underlying population showed relatively homogeneous risks of MND without distinct differences across regions in Scotland despite wide socioeconomic inequalities, large differences in urban/rurality and housing conditions in the Scottish population at risk.

Estimated lifetime risk of MND modelled by the fitted logistic curves agrees with previously published estimates^48^ and can be interpreted as the risk of being diagnosed with MND over the typical (expected or mean) number of years lived for each sex in the UK. However, it is apparent that the lines after this point are diverging and, that particularly for males that live longer, the risk may be substantially higher than this. The asymptote of the fitted curve may be interpreted as the proportion of people who would be expected to develop MND based on these trends if it were not for some other competing risk of death.

Previous studies examining the association between SES and risk of MND have shown inconsistent findings, both in whether a link exists and the direction of the effect. In our study, before adjusting for age and sex, a significant higher risk of MND was found in the areas with the highest SES compared to those that were the most deprived. As initially intriguing as this seems however, the association was nullified and lost significance after adjusting for age and sex. Our study is in agreement with others, where no association was identified between SES and risk of MND.^27,29^ A different approach to this question was adopted by Rooney et al, who found no difference between the indices of social deprivation in the areas within the identified clusters and the rest of the country.^26^ Among those that found associations, two studies used income^21^ and education^22^ as indicators of SES, and showed that higher SES was associated with significantly higher risk of ALS. Another study^23^ found a positive correlation between provincial gross domestic product per capita and mortality of ALS in Spain. In contrast, Palo et al.^25^ found lower SES was associated with higher risk of MND. While most studies examined SES around the time of diagnosis, Martyn et al.^24^ and Chancellor et al.^28^ examined several factors reflecting socioeconomic deprivation in childhood and the risk of developing MND later in life,^24^ they too found some contradictory results. It is worth noting that comparison between these studies requires cautious interpretation due to several methodological issues, including different definitions and exposure windows of SES, individual or areal levels of SES, study designs, confounding adjustment and sample sizes.

In summary, our results suggest there is insufficient evidence and no clearly plausible hypothesis to support a link between SES and the risk of MND. Furthermore, although it may be that such an association does exist in some locations, it should be noted that without adequate data to control for small-area variations in age- and sex-structure of the population at risk, these nuisance variables are liable to cause spurious associations. Modest inaccuracies in denominators may plausibly explain any modest effect-sized association and may be difficult to detect. A lack of association with SES is an important finding as many modifiable lifestyle factors are associated with SES, and so these may be pertinent null associations in that they make strong contributions to missing environmental MND risk from any risk factor that is associated with SES unlikely unless counter-acted by other missing factors.

Contrary to several previous studies,^12–15^ we did not identify any significant population cluster or distinct risk patterns of MND using a variety of statistical and methodological approaches, although lower SIRs of MND were found in two health boards (Highland and Lanarkshire). Given the high case ascertainment rate of the CARE-MND,^30^ it is unlikely that under-ascertainment led to lower SIRs. Also, when more granular DZs were used to geocode cases, no significant cluster of lower risk was identified in these two regions. One possibility may be that this is an artefact of migration in the pre-diagnosis period due to symptoms of the disease. Patients who lived in Lanarkshire and Highland might move closer to tertiary neurological centres during the period of diagnostic uncertainty prior to diagnosis. This may also partially explain why Greater Glasgow and Clyde, a major tertiary referral centre, had a trend towards higher observed cases. Similarly, while higher SIR was shown in Dumfries and Galloway, no significant cluster of high risk was detected in the region. However, care for MND in Scotland is community focussed and migration for access, therefore, is thought to be uncommon.

In addition, the small but significant result of the global Moran’s I indicated that there might be some small amount of spatial non-randomness (autocorrelation) in MND distribution at a local level. Thus, it remains possible that one or more of the non-significant clusters identified are in fact real or that some clustering exists in these data below the level at which our tests are powered to detect. We viewed it particularly worth investigating these possible clusters as the power of cluster detection methods, though influenced by many factors, is generally lower where the disease is rare,^45^ and there are contributions from clusters (e.g. due to a local environmental risk factor) mixed in amongst spatially random cases. In addition, our power to detect clusters may be diminished if location of residence at the time of diagnosis is not the best measure of a person’s geographic risk. For example, clusters based on location of workplace, schooling or within families, may identify novel environmental or genetic risk factors. Thus, we reviewed the cases within possible clusters but did not identify epidemiological links between the cases. Consequently, future surveillance is intended to prospectively monitor rates in NHS Highland, Lanarkshire and around the town of Tullibody.

The subset of causes that lead to spatiotemporal variation (i.e. clusters) are particularly valuable to identify as these may be the most modifiable of the causal factors. Nevertheless, a key limitation of the spatial approach is that the causal genes or environmental factors can only be identified if they are themselves are sufficiently spatially structured and exposure over time or space varies at a relevant scale. Therefore, absence of the detection of clusters or spatial stratification/association is not an argument for the unimportance of gene variants or environmental factors but rather suggests that these factors are either ubiquitous or sporadically but randomly distributed themselves over the study area. This information may still be helpful in identifying such risk factors.

A major strength of this study is the comprehensive depth of detail and capture of MND cases across Scotland enabled by CARE-MND. Since the registry has been shown to capture 99% of the MND cases in Scotland,^30^ it is unlikely that ascertainment bias would occur in our study. In addition, the large sample size has allowed us to have statistical power to detect whether spatial clusters existed in Scotland. Another strength is that the DZs used in this study covered small, yet stable areas and population for both Poisson regression and cluster analysis and that age- and sex-specific population estimates are available at this level. The DZ was also useful in its direct linkage with SIMD. Furthermore, SIMD was designed to comprise seven key domains of social deprivation, including income, employment, education, housing, health, crime, and geographical access. Since these indicators should not be used interchangeably,^49^ a composite index like SIMD might be a more appropriate and comprehensive indicator of SES than a single measure.

Nevertheless, our study is not without its limitations. First, complete genetic information is not available in the current database. Genetic risk factors and the interactions between genetic and environmental risk factors have been suggested as explanations for the geographic variations and clusters of MND.^14,15^ Furthermore, only residence at diagnosis rather than lifetime residence was available in the database. It is possible that patients had been exposed to risk factors many years before disease onset. However, without lifetime residential history, it was impossible to assess different incubation periods or potential exposure at different time points. Nevertheless, given that there is no well-established hypothesis regarding environmental risk factors or the time required from exposure to disease onset, residence at diagnosis still provides useful information about how cases are distributed throughout Scotland.

For future studies in search for geographical or socioeconomical clustering of MND, genetic data would be important in determining the association between the spatial distribution of genes and MND cases. In addition, whilst MND might seem a rare disorder in terms of incidence, considering the lifetime risk estimated in our study, healthcare resources and research in MND are needed. The findings in our study also provide evidence for the implication of distributing healthcare resources. Specifically, no difference in MND risk between rural and urban areas was found. Scotland has invested in community-led MND care, and therefore our data suggest that there is no great trend towards moving in the pre-diagnostic period or differential ascertainment of cases between urban and rural resident individuals. This may underlie the importance of investment in community and mobile health services, which provide better access for those in need. Regular cluster analysis using data from national registers would also be useful in allocating resources of healthcare and research regarding MND.

### Conclusion

This is the first study reporting how SES and geography affect risk of MND using Scottish population-based data. Our study did not identify an association between SES and MND risk after adjusting for age and sex. In addition, by using two approaches, including SaTScan and Bayesian modelling, we did not find evidence for spatial clusters or regional excess risk of MND throughout Scotland. Our finding of a small but significant amount of spatial autocorrelation would, however, be consistent with some small clusters existing on a background of most cases being randomly distributed.

## Supporting information

STROBE

