## Supplement for "Motor Neuron Disease is not clustered or associated with socioeconomic deprivation in Scotland"

**Supplemental material**

**eMethod. Cluster detection by Satscan**

Scanning was performed separately for spatial clusters of higher or lower than expected rates, by examining circular windows with continuously varying sizes, adjusting the expected number of cases for the age and sex of the underlying population. Areas were judged to fall within the scanning window if their population weighted centroid did, and a discrete Poisson model was applied to fit the underlying distribution. The maximum circular window size was set to be 50% of the population at risk. Although there is no established hypothesis of MND cluster size, the maximum reporting size of clusters were set to be 30km based on the findings of previous literature of relatively small, local clusters as well as to enhance interpretability and practical actions (outbreak investigation) upon detection of clusters. Monte Carlo simulations, and the extreme Gumbell distribution, were used to obtain pseudo-p values for the most likely clusters detected.

**eFigure 1. Bayesian smoothed risk map, stratified on gender, adjusting for age of underlying population**

A. Female

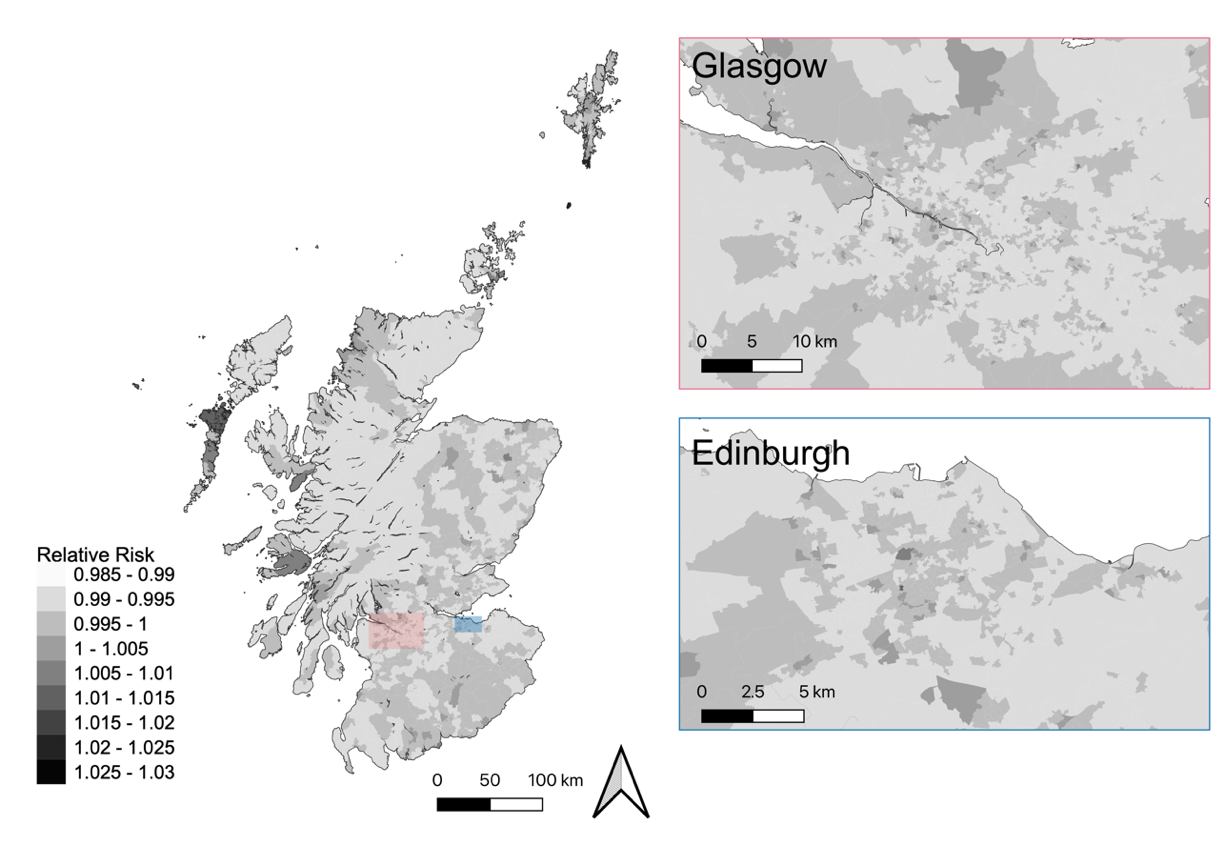

B. Male

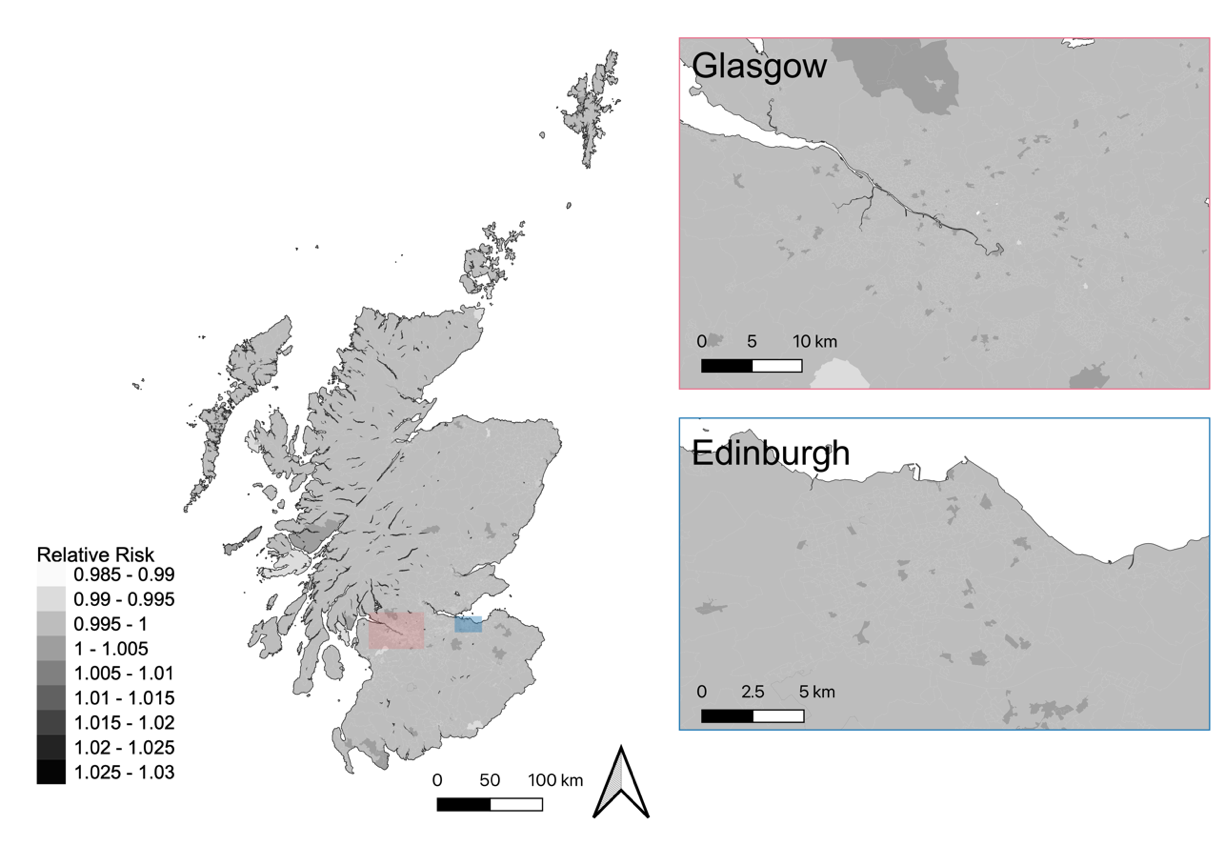

**eFigure 2. Bayesian smoothed risk map, stratified on age at diagnosis (≥ 55 years and < 55 years), adjusting for age and sex of underlying population**

A. Age at diagnosis **≥ 55 years**

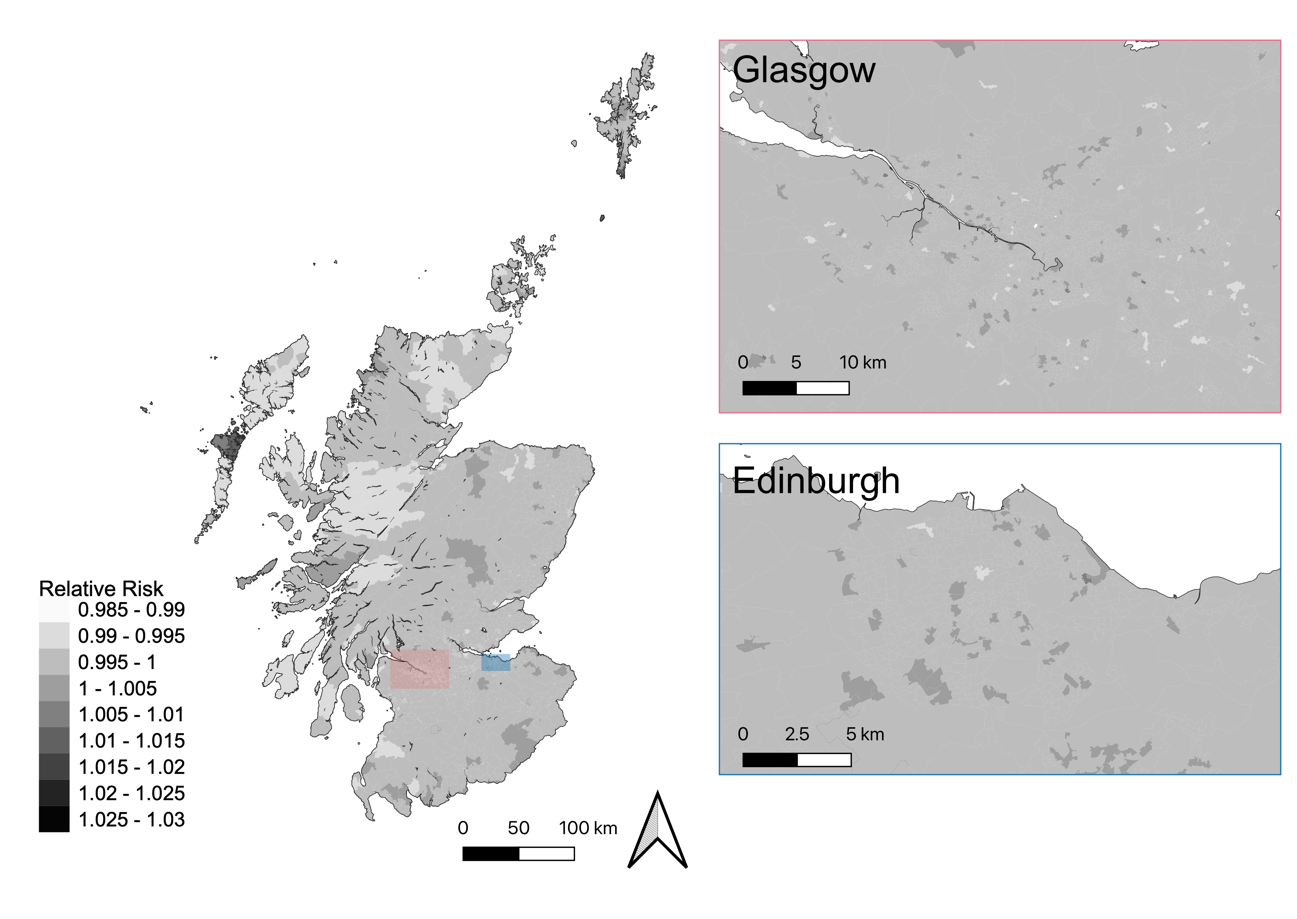

B. Age at diagnosis **< 55 years**

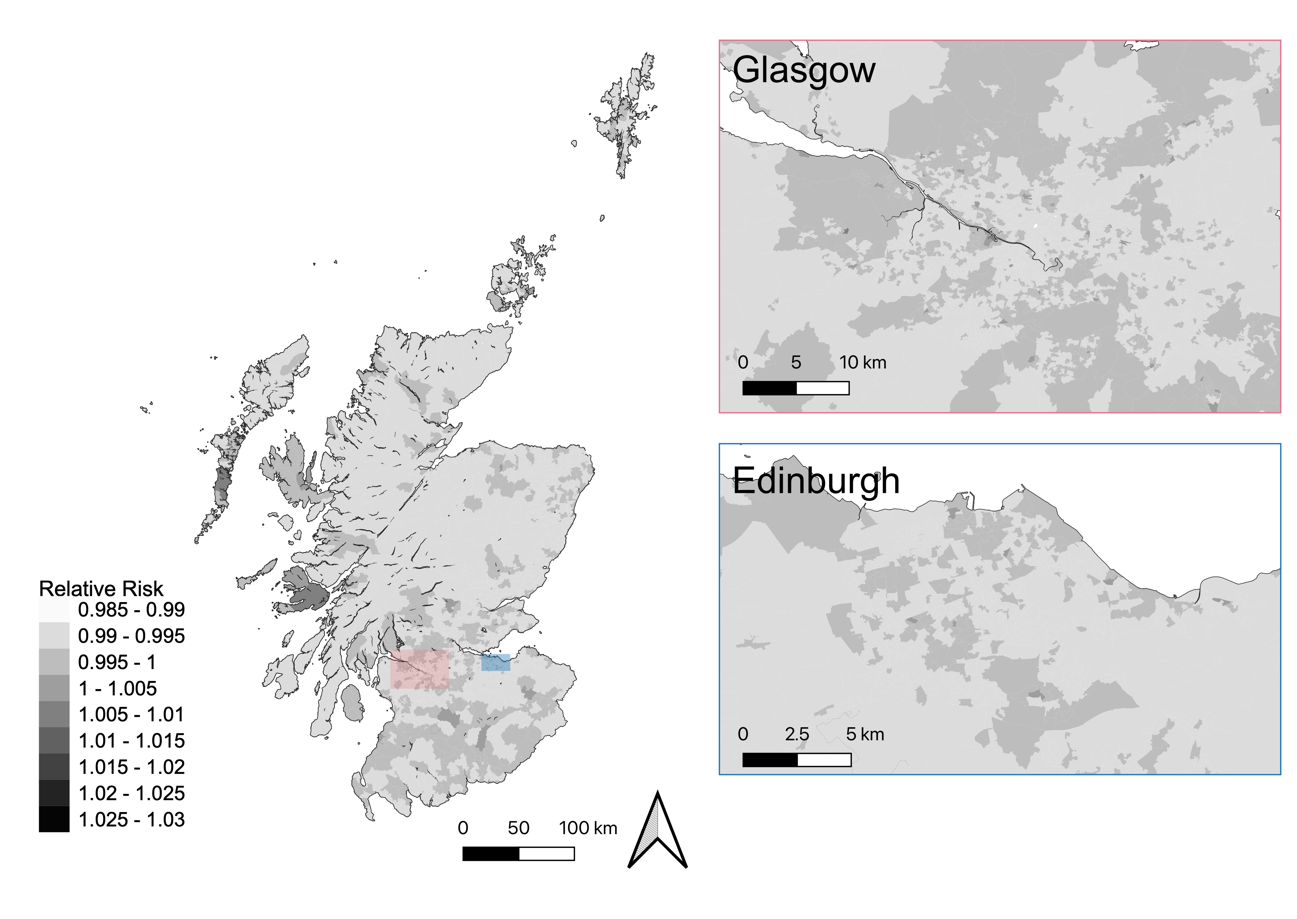

**eFigure 3. Bayesian smoothed risk map, stratified on classification of MND, adjusting for age and sex of underlying population**

A. Amyotrophic lateral sclerosis (*n*=807)

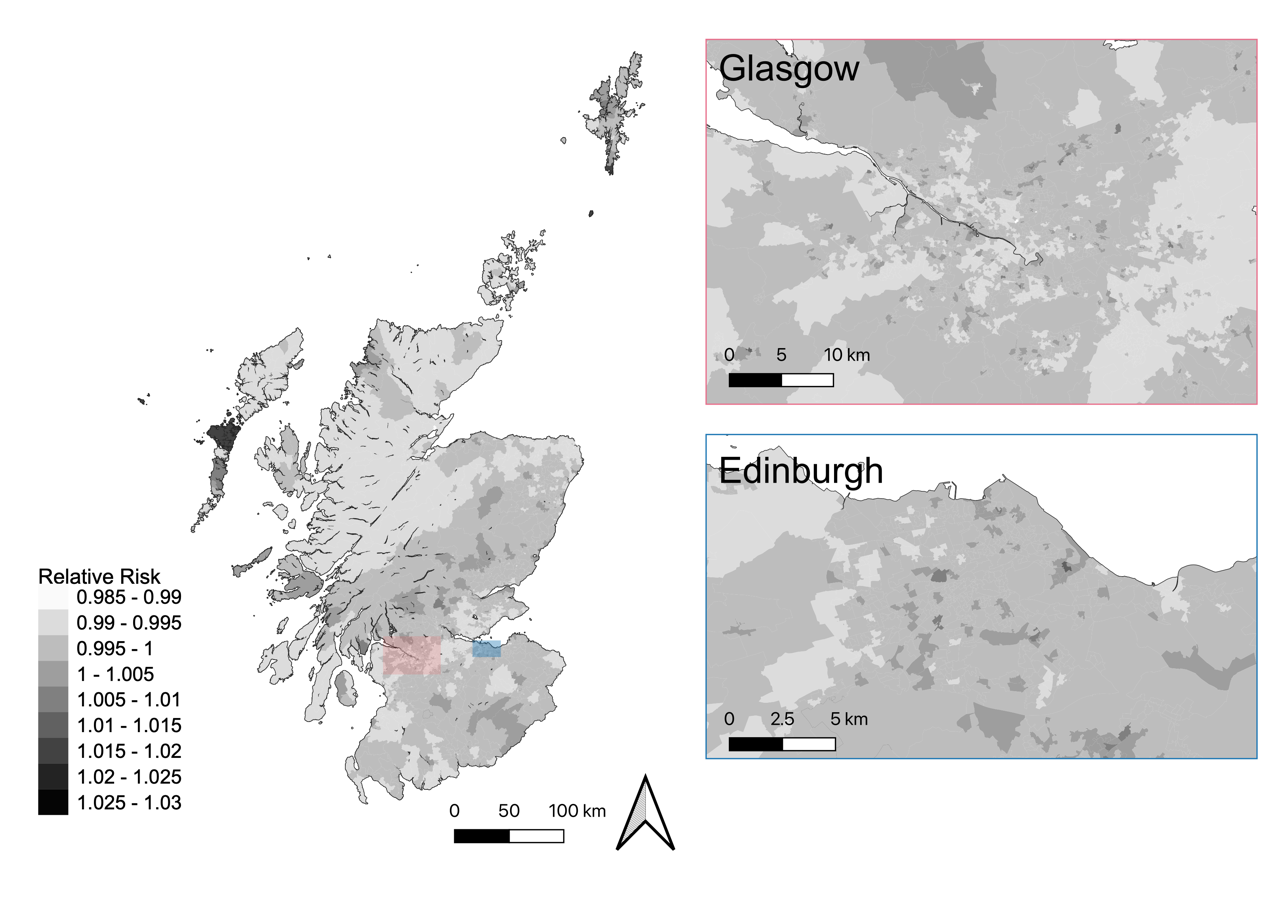

B. Progressive bulbar palsy (*n*=104)

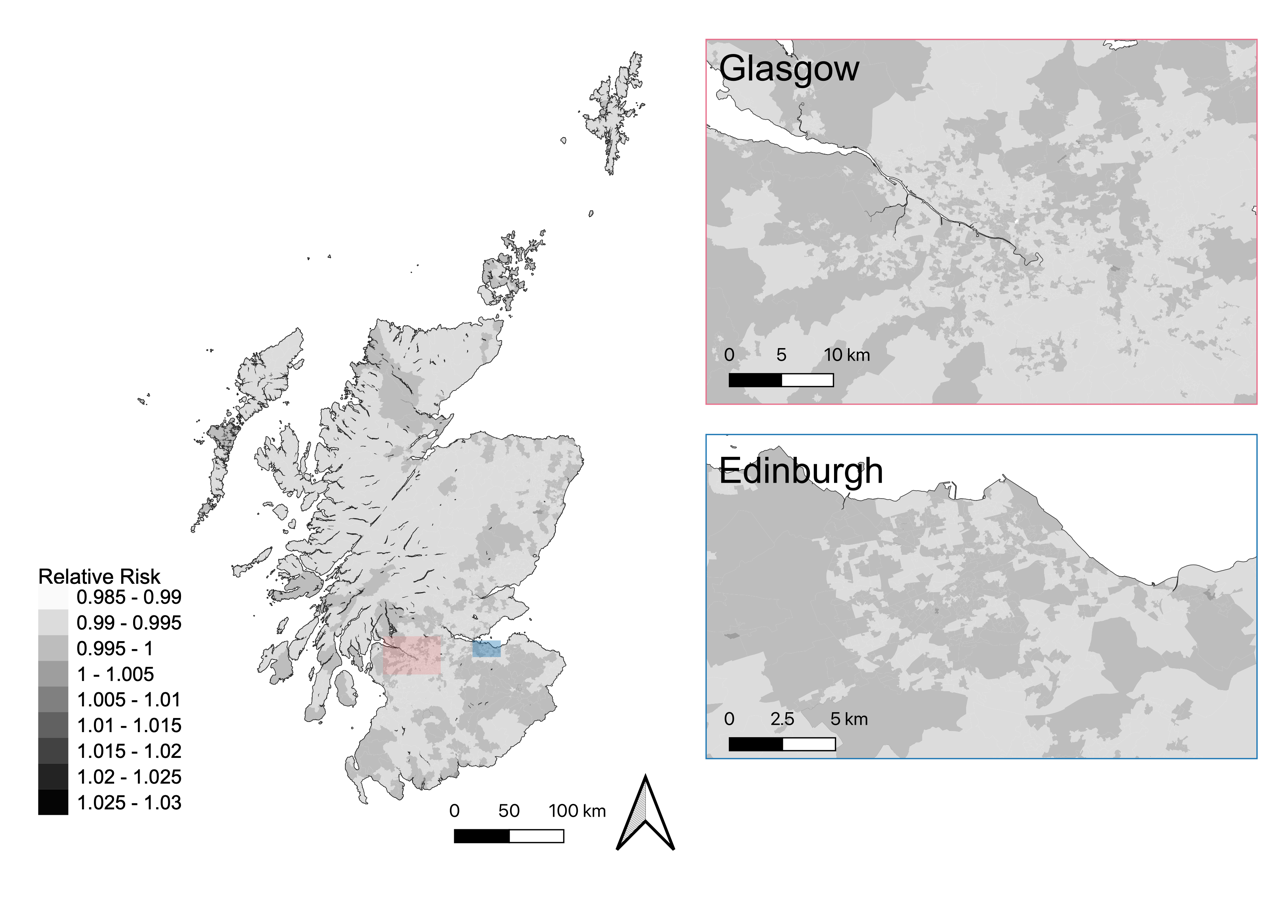

C. Motor neuron disease with frontotemporal dementia (*n*=70)

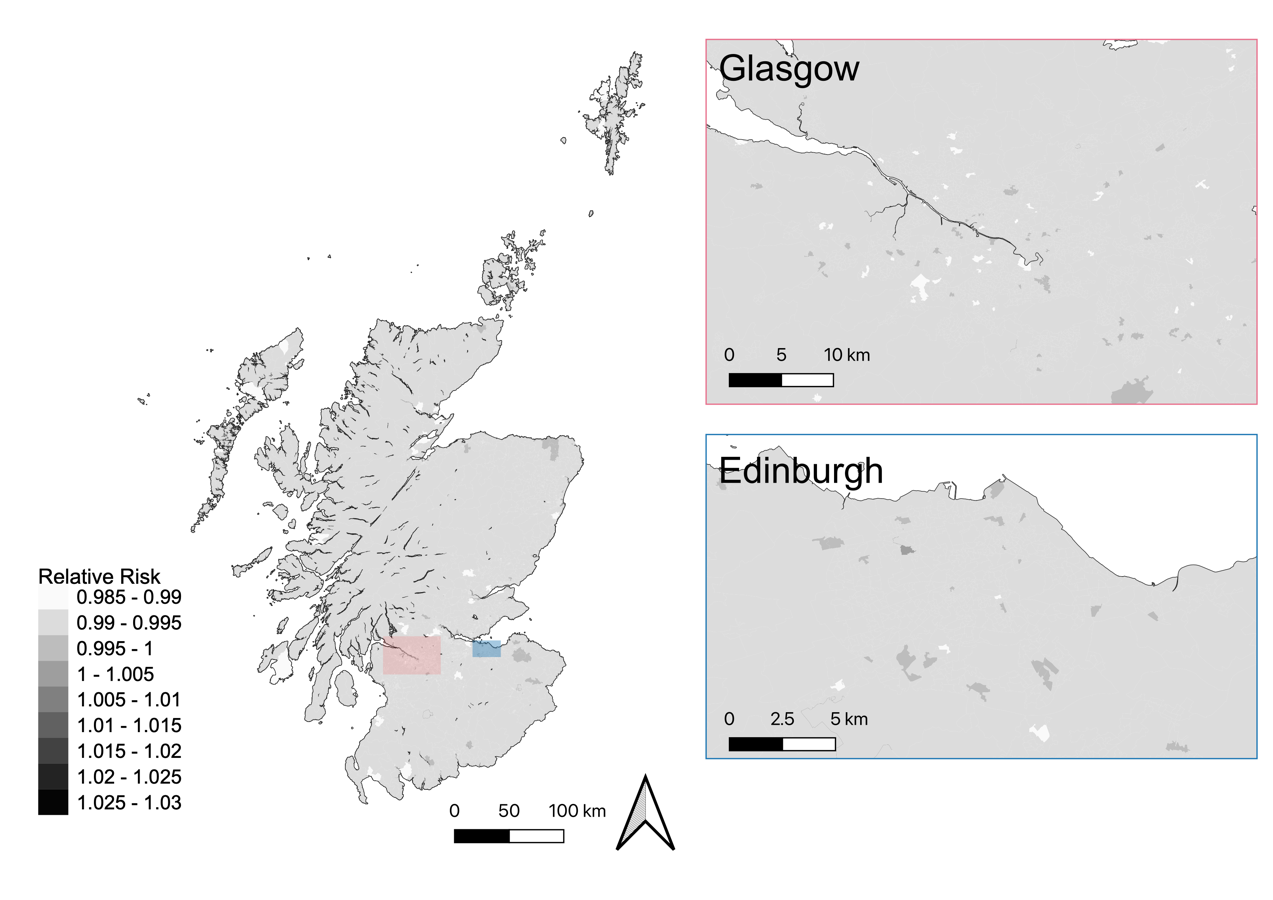

**eFigure 4. Unsmoothed risk map of motor neuron disease in Scotland.** (Note: The scale of legend is different from other maps due to the wide range of relative risks before Bayesian smoothing)

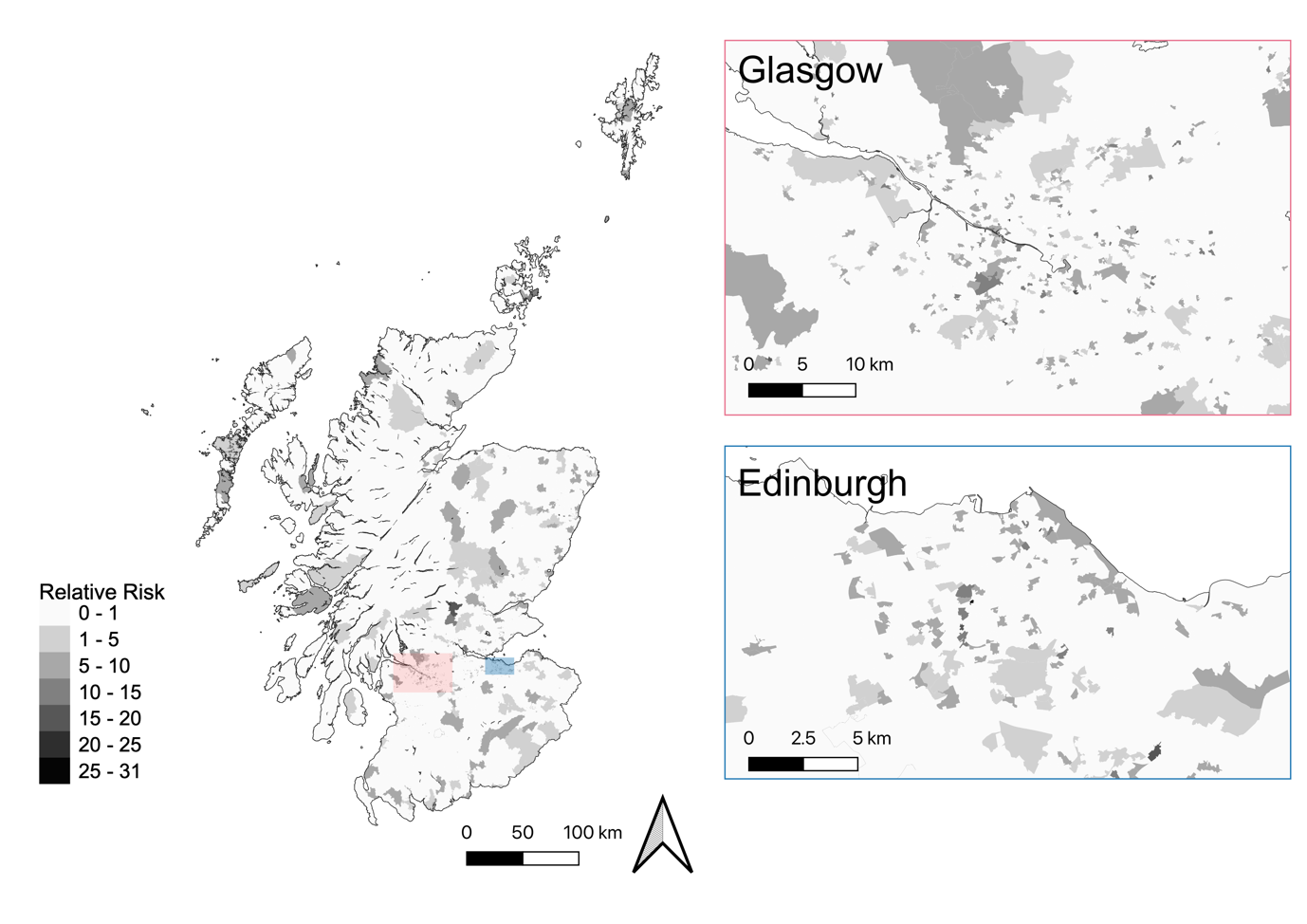

**eFigure 5. Scan for low-risk clusters of motor neuron disease in Scotland.** (maximum scanning window 50% population at risk, maximum reporting size 30km) None of them reached statistical significance.

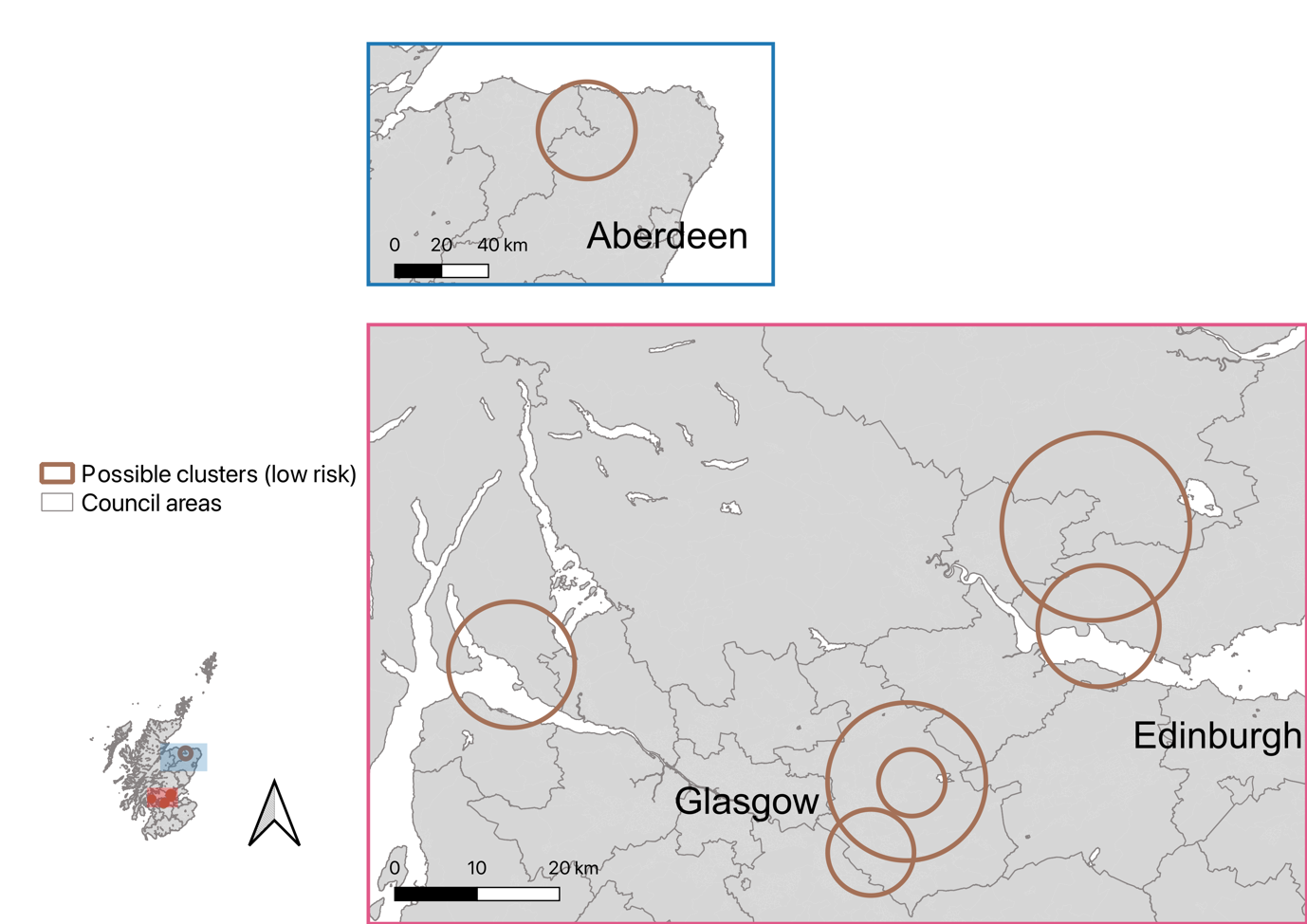

5

2

6

3

1

4

7

| **Cluster number (see above)** | **Size (radius)** | **Observed cases** | **Expected cases** | **Average population** | **Relative risk** | ***p*** |
| --- | --- | --- | --- | --- | --- | --- |
| 1 | 5.22 km | 9 | 26.56 | 136534 | 0.33 | 0.58 |
| 2 | 11.38 km | 0 | 6.96 | 28372 | 0 | 0.85 |
| 3 | 20.75 km | 1 | 10.19 | 41914 | 0.097 | 0.90 |
| 4 | 9.58 km | 18 | 38.31 | 198669 | 0.46 | 0.90 |
| 5 | 7.36 km | 1 | 10.16 | 44820 | 0.098 | 0.90 |
| 6 | 7.64 km | 3 | 13.92 | 58589 | 0.21 | 0.98 |
| 7 | 4.04 km | 0 | 6.28 | 33253 | 0 | 0.99 |

**eFigure 6. Scan for high-risk clusters of motor neuron disease in Scotland.** (maximum scanning window 50% population at risk)

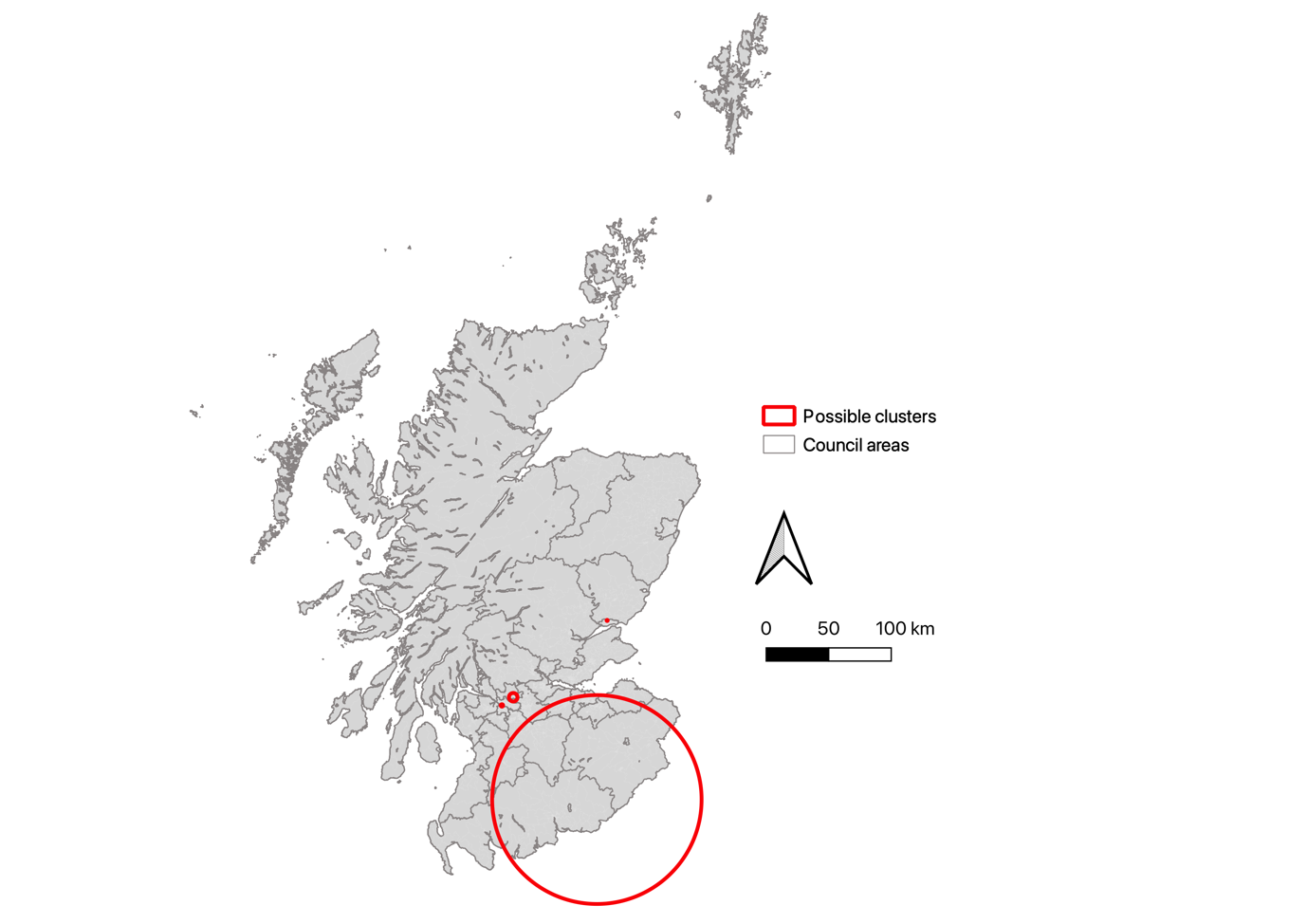

3

4

2

1

| **Cluster number (see above)** | **Size (radius)** | **Observed cases** | **Expected cases** | **Average population** | **Relative risk** | ***p*** |
| --- | --- | --- | --- | --- | --- | --- |
| 1 | 0.97 km | 11 | 2.31 | 17542 | 4.79 | 0.455 |
| 2 | 0.44 km | 4 | 0.25 | 1346 | 16.06 | 0.827 |
| 3 | 83.53 km | 241 | 191.87 | 928619 | 1.33 | 0.855 |
| 4 | 3.44 km | 23 | 9.55 | 42807 | 2.44 | 0.964 |

Note: Clusters 1, 2 & 4 are the same as the clusters identified in the scan with maximum scanning window 50% population at risk and maximum reporting size 30km (Figure 2).

**eFigure 7. Scan for high-risk clusters of motor neuron disease in Scotland.** (maximum scanning window 50% population at risk, elliptic windows)

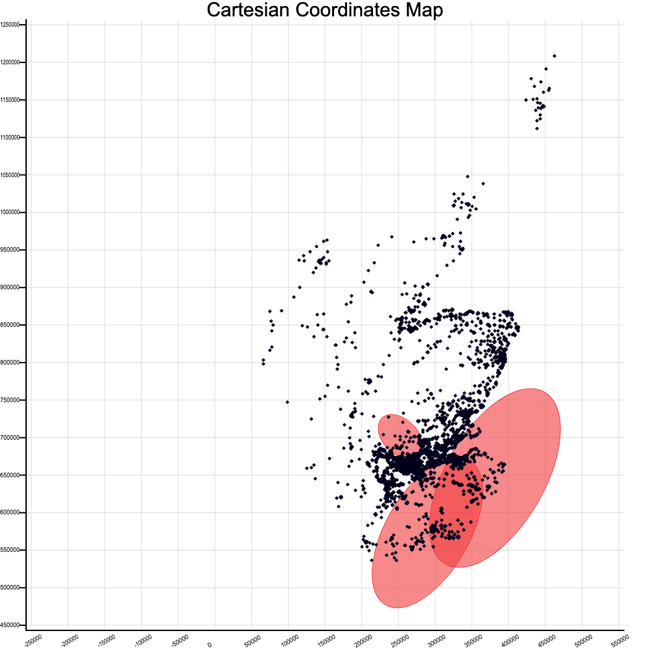

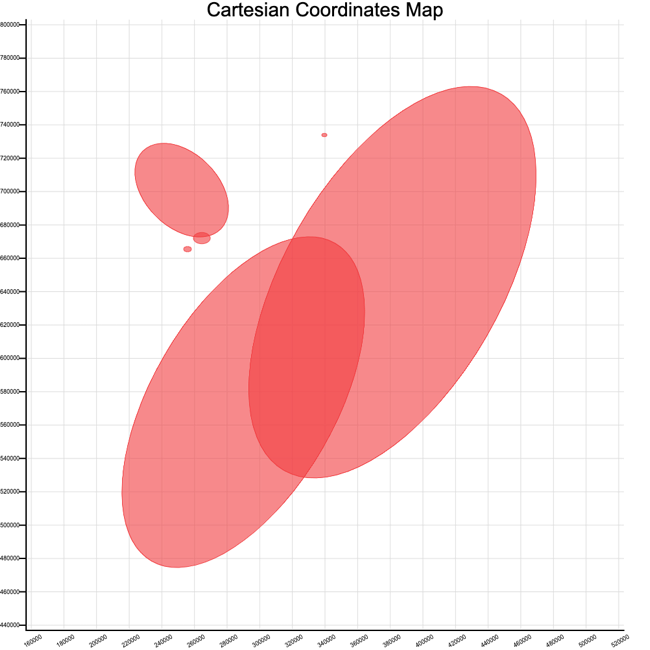

4

3

6

2

5

1

Glasgow

Edinburgh

Glasgow

Edinburgh

| **Cluster number (see above)** | **Observed cases** | **Expected cases** | **Average population** | **Relative risk** | ***p*** |
| --- | --- | --- | --- | --- | --- |
| 1 | 14 | 2.71 | 24750 | 5.23 | 0.076 |
| 2 | 209 | 157.64 | 763122 | 1.4 | 0.718 |
| 3 | 5 | 0.39 | 2321 | 12.98 | 0.836 |
| 4 | 24 | 9.48 | 42779 | 2.57 | 0.900 |
| 5 | 214 | 167.44 | 842243 | 1.34 | 0.995 |
| 6 | 40 | 21.20 | 96261 | 1.92 | 0.998 |

**eAppendix. Summary of possible clusters based on CARE-MND data**

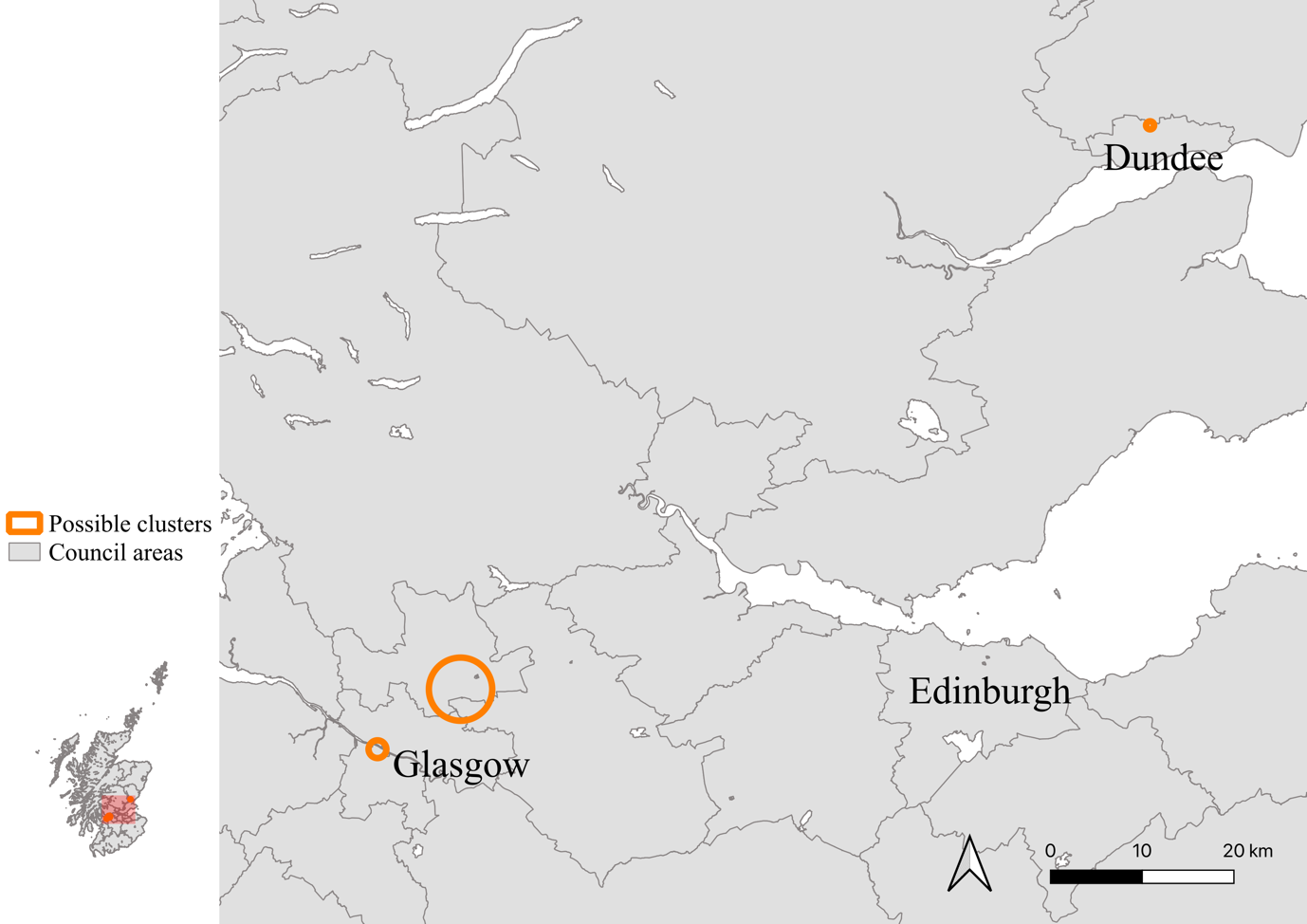

Possible cluster 1 – Glasgow Clyde River

This possible cluster in Glasgow City centre contains 11 MND cases located both north and south of the Clyde River, within the areas of Govan, Thornwood and Partick. These cases were all diagnosed between 02/2015 and 09/2020 and are all currently deceased. Only one of the cases in this cluster had a family history of MND. In terms of social history, there is a large variation in socioeconomic status within these areas, with Govan being in the 1^st^ most deprived SIMD decile, in contrast to Thornwood, which is in the 10^th^ least deprived SIMD decile. The occupational history of the cases is as follows: 2 domestics, 1 chartered surveyor, 1 factory worker, 1 office worker, 1 fraud investigating officer, 1 police officer and 4 cases in which the occupation was not known. Overall, given the lack of any obvious connection between the cases and the geographical barrier created by the Clyde River, it is likely that this possible cluster arose by chance.

Possible cluster 2 – Dundee (Downfield area)

This possible cluster contains 4 cases located in the Downfield area of North Dundee. These cases were diagnosed between 01/2015 and 01/2019. As of July 2021, 2 of the cases are alive and 2 are deceased. There is no family history of MND in 3 of the cases, but we do not have data for the last case. In terms of occupations, 1 person worked as a joiner and another as a student nurse. Unfortunately, there is no occupational history data available for the 2 deceased cases. Overall, we have found no connections between the cases, however these could benefit from further investigation to obtain some of the missing data.

Possible cluster 3 – Glasgow North (Bishopbriggs and Kirkintilloch)

This is the largest possible cluster identified, containing 23 cases located in the towns of Bishopbriggs and Kirkintilloch, just north of Glasgow City. Out of the 23 cases, 4 are alive and 19 are deceased as of July 2021. All cases were diagnosed between 01/2015 and 08/2020. 2 of the cases had a family history of MND, 18 had no family history and data was missing for 3 of the cases. Past occupational history was very diverse, including: 1 psychologist, 4 office workers, 1 police officer, 1 lift engineer, 1 joiner, 1 nurse, 1 minister, 1 factory worker, 2 teachers and 1 domestic. Occupational history was missing for 9 of the cases. Overall, we have found no connections between the cases, however these cases could benefit from further investigation to obtain some of the missing data.
